# KNOWLEDGE, ATTITUDES AND PRACTICE AMONG HEALTHCARE WORKERS TOWARDS COVID-19 PREVENTIVE MEASURES AT WOMEN AND NEW-BORN HOSPITAL, UNIVERSITY TEACHING HOSPITAL, LUSAKA, ZAMBIA

**DOI:** 10.1101/2021.12.20.21267829

**Authors:** Joyce Nambela Shampile, Godfrey Lingenda, Mowa Zambwe, Peter J. Chipimo

## Abstract

**Objective:** To assess the knowledge, attitude and Practices among healthcare workers towards covid-19 preventive measures at Women and New-born Hospital of the University Teaching Hospitals in Lusaka.

**Methods:** Cross-sectional study conducted at WNH-UTH, Lusaka. Convenient sample of 264 frontline healthcare workers responded to self-administered questionnaire to determine their knowledge, attitudes and practices on COVID-19 preventive measures.

**Results:** Majority (31.9%) of the respondents were aged 25 – 29 years. The study revealed that 63.4% had a good knowledge, 60.3% had positive attitudes and 59.9% had a good practice. Attitude was positively related with practice (*r* = 0.524, p < 0.001) and knowledge (*r* = 0.469; p < 0.001). Further, knowledge was positively correlated with practice (*r* = 0.51; p < 0.001). Bivariate analysis results showed that only high knowledge score (75.6%; p < 0.001) and high attitude score (77.6%; p < 0.001) was associated with an increase in good practice among healthcare workers towards Covid-19 preventive measures.

**Conclusion:** The study showed the need for continued assessment of Knowledge Attitude and Practice among healthcare workers towards Covid-19 preventive measures. It further showed the need of designing interventions aimed at encouraging sustained compliance to preventive measures among healthcare workers to prevent COVID-19 transmission.

**ARTICLE SUMMARY:** *Strengths and Limitations:* This study was conducted at Women and Newborn Hospital of the University of Zambia and described the Knowledge, Attitude and Practice among Healthcare workers towards Covid-109 preventive measures. The study showed the need for continued assessment of Knowledge Attitude and Practice among healthcare workers towards Covid-19 preventive measures. This study has the potential to yield significant benefits for the participants and the community at large. The study is reproducible and feasible with results which can be used in designing interventions aimed at encouraging use of preventive measures available to healthcare workers to prevent COVID-19 transmission. The study was a cross-sectional study which limits our ability to make causal inferences. Further the study was conducted at only one hospital which is located in the capital city and so the findings cannot be generalized to HCW in other parts of Zambia.

## Introduction

Coronavirus disease 2019 (COVID-19) was first reported in Wuhan City of China in December 2019. From that time on, the epidemic has spread to over 223 countries and has been declared a global pandemic by the World Health Organization^1^. The pandemic has grown to more than 100 million positive COVID-19 cases, with at least 2.4 million deaths globally^1^. In Zambia, from the first two cases first recorded in March 2020 and since then, more than 69, 437 confirmed cases with 951 deaths have been recorded.^1,2^

Frontline healthcare workers (F-HCWs) are prone to getting infected^3^. Global reports have indicated a significant number of COVID-19 deaths among health workers globally. For example, it was reported in early September 2020 that the number of COVID-19 deaths among health workers worldwide was at least 7000.^3^ The countries that were estimated to have the most deaths of health workers were Mexico (1,320), the United States of America (1,077) and the United Kingdom (649) ^3^ A study conducted in the United Kingdom as well as in the United States found that frontline healthcare workers are more likely to test positive for COVID-19 than the general community ^3^. Inpatient settings appeared to pose the most risk followed by nursing homes, and outpatient hospital and clinics ^4^

Negligence and lack of knowledge have been identified as major causes of a prevailing rate and transmission of COVID-19 infection among local F-HCWs at the initial stage of the disease outbreak in places such as Wuhan and Henan. However, the infection rate was noted to reduce among the F-HCWs after a significant increase in knowledge about COVID-19 thus enhancing compliance to preventive measures.^5,6^Limited number of studies have been conducted in Zambia to assess the knowledge, attitudes and practices of frontline healthcare workers. The only study we were able to find investigated KAP only among specific Health-worker categories (Laboratory staff) but did not include other health workers involved in the fight against COVID-19 pandemic in the country. ^7^

Thus, the aim of this study is to assess the knowledge levels, attitudes and practices among healthcare workers towards covid-19 preventive measures at Women and New-born Hospital of the University Teaching Hospitals.

## Methodology

This was a cross-sectional study design which enrolled 266 healthcare worker. The study population included healthcare workers, aged 18years and over, who had been working at the Women and Newborn Hospital from the time of the COVID-19 pandemic and agreed to participate in the study.

The study was questionnaire based where data on demographic factors, knowledge, attitudes and practices on COVID-19 prevention among healthcare workers was collected using a semi-structured and self-administered questionnaire. The questionnaire was hand delivered and contained a specific number of questions on the socio-demographic characteristics, knowledge, attitude and practices on COVID-19 among healthcare workers.

The data was analyzed using Stata v.14.0 and included, descriptive statistics presented as summary measures and tables. Chi-Square was used to measure association and difference among demographic variables. Clustered bar charts were used to compare data across categorical variables. The relationship between knowledge, attitude, and practice was determined using Spearman’s rho correlation test. Finally, knowledge, attitudes and practice were modelled using multiple logistic regression model to identify predictors of sufficient knowledge, positive attitudes and good practice among healthcare workers. This model was built using bivariate analysis which was performed to identify predictors associated with the practice. All predictors with a p-value < 0.25 were assessed for inclusion in the model using the likelihood ratio tests. All variables which improved the model (p value < 0.05) were retained in the final model.

## Results

The study had 264 healthcare workers of whom majority (31.9%) were aged 25 – 29 years and only6.5% aged above 45 years of age. About40.6% were married while53.3% were single. Over 70% of the healthcare workers had received training on COVID-19 while 29.2% had not. Additionally, 67.2% had college education while 19.8% had attained a University education. Over half of the participants (63.1%) were nurses, while just over 10% (12.9%) were general workers.

### Correlation: Knowledge, Attitudes and Practice

Table 2 shows the Spearman’s rank correlation matrix for Knowledge, Attitude and practice. The results revealed medium strength positive correlation which were all significant at 99% confidence interval. The correlation coefficient was stronger for Practice and Attitude (r=0.524) Practice and Knowledge (r=0.510) vs. Knowledge and Attitude (r=0.469)

**Table 1:** Socio-demographic Profile of Healthcare Workers at Women and New-born Hospital

| **Characteristic** | **n(%)** |
| --- | --- |
| Age group |  |
| 20 - 24 | 39 (15) |
| 25 - 29 | 83 (31.9) |
| 30 - 34 | 38 (14.6) |
| 35 - 39 | 48 (18.5) |
| 40 - 45 | 35 (13.5) |
| Above 45 years | 17 (6.5) |
| Marital Status |  |
| Single | 139 (53.3) |
| Married | 106 (40.6) |
| Separated | 5 (1.9) |
| Divorced | 4 (1.5) |
| Widow | 7 (2.7) |
| Training on COVID-19 |  |
| No | 73 (29.2) |
| Yes | 177 (70.8) |
| Highest Level of Education |  |
| Primary | 11 (4.2) |
| Secondary | 23 (8.8) |
| College | 176 (67.2) |
| University | 52 (19.8) |
| Profession |  |
| Nurse | 166 (63.1) |
| Doctor | 9 (3.4) |
| General worker | 34 (12.9) |
| Laboratory Technologist | 15 (5.7) |
| Radiographer | 5 (1.9) |
| Other | 34 (12.9) |

**Table 2:** Correlation: Knowledge Attitude and Practice.

| Variable (Median ± IQR*) | Attitude | Knowledge | Practice |
| --- | --- | --- | --- |
| Attitude (16 ± 3) | 1 | | |
| Knowledge (21 ± 4) | 0.469** | 1 | |
| Practice (20 ± 5) | 0.524** | 0.51** | 1 |

\*IQR = Interquartile range
\*\* $p$ -value = <0.001

**Table 2:** Bivariate association between Knowledge, Attitude and Practice.

| Characteristics |  | Practice on COVID-19 |  |  |
| --- | --- | --- | --- | --- |
|  |  | Poor (n = 101) | Good (n = 151) | p-value |
| Attitude | Negative (n = 100) | 67(67%) | 33(33%) | <0.001 |
|  | Positive (n = 152) | 34 (22.4%) | 118 (77.6%) |  |
| Knowledge | Poor (n = 90) | 61 (67.8%) | 29 (32.2%) | <0.001 |
|  | Good (n = 156) | 38 (24.4%) | 118 (75.6%) |  |

## Knowledge, Attitude and Practice Scores

To determine the Knowledge, Practice and Attitude, the summative scores of each was converted to a binary variable using the mean as the cut-off point, these being (good or poor), attitude (negative or positive) and practice (good or poor) respectively. The mean cut-off point for attitude was 76% (15/20 questions), 80% (19/24) questions for knowledge and 72% (18/25 questions) for practice. The study revealed that 63.4% had a good knowledge, 60.3% had positive attitudes and 59.9% had a good practice.

**Figure 1:**
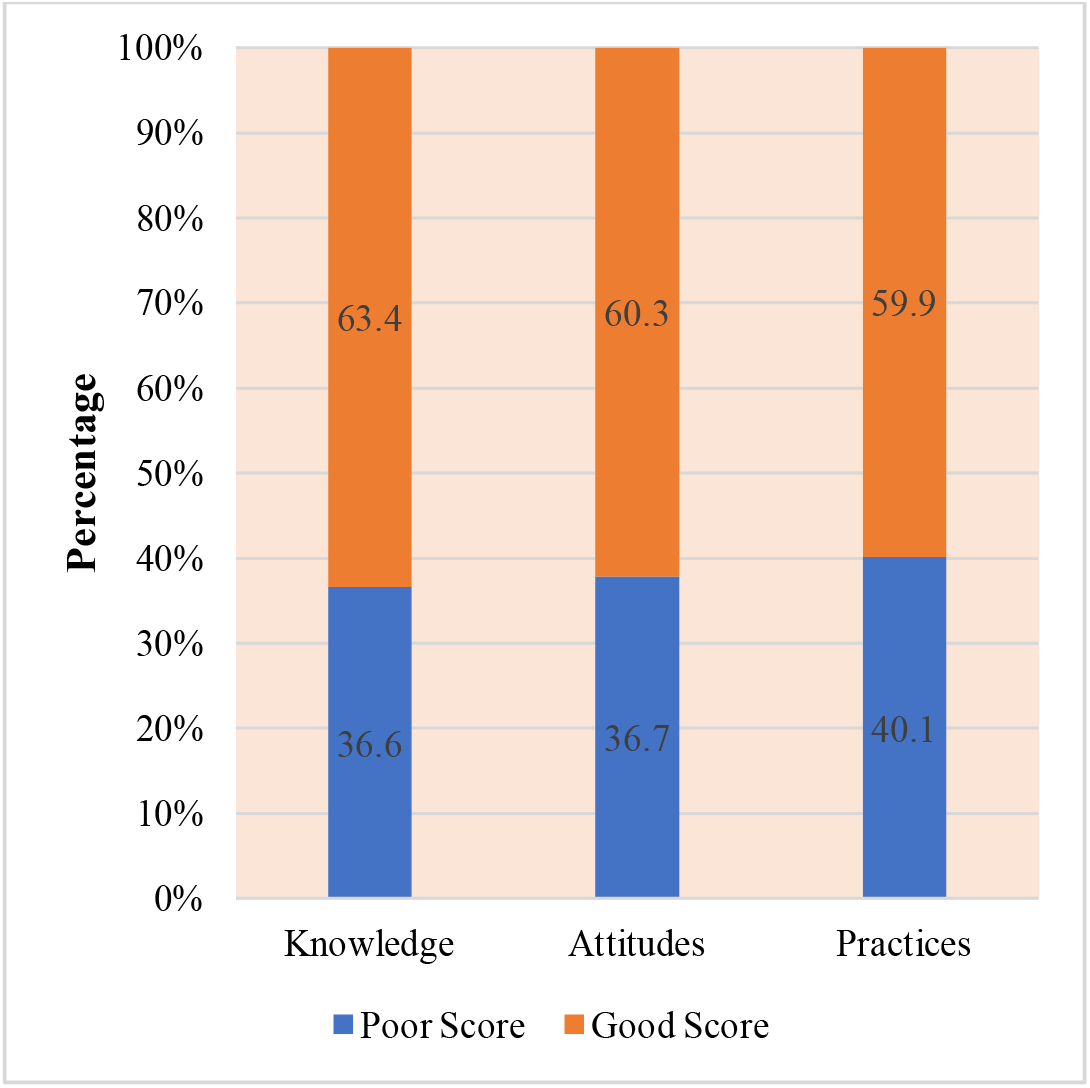
Knowledge, Attitude and Practice Scores among Healthcare Workers at Women and Newborn Hospital.

Table 3 shows the bivariate relationship between Attitude, Knowledge and Practice. It was observed that healthcare workers with a positive attitude towards COVID-19 prevention were more likely to have good practices (p < 0.001) and so are the healthcare workers with good level of knowledge (p < 0.001). Further the results show that a good score on knowledge also influenced attitudes towards COVID-19 prevention among healthcare workers (p < 0.001).

### Source of Information on COVID-19 preventive measures

Most of the participants (84.1%) indicated that their main source of information was via TV followed by healthcare workers (76.1%), radio (63.6%), friends (59.8%), newspapers (NPs) (54.9%) and finally other sources (4.5%).

**Figure 2:**
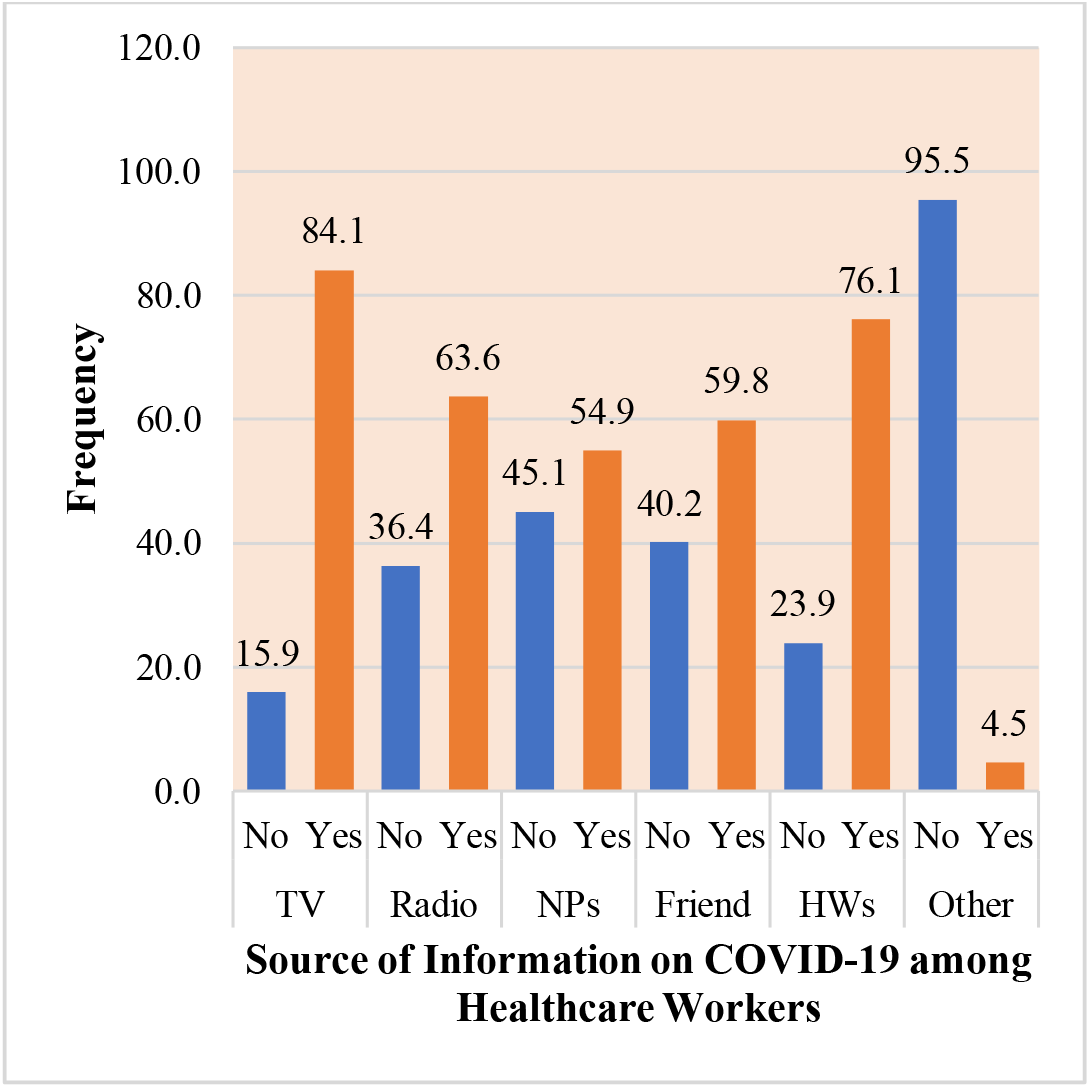
Source of Information on COVID-19 preventive measures among Healthcare Workers at Women and Newborn Hospital.

## Discussion

The study revealed that most of the healthcare workers had sufficient knowledge on COVID-19 and its prevention. The study showed that 63.4% of the healthcare workers had sufficient knowledge on COVID-19 prevention. In other words, 63.4% of healthcare workers scored over 80% (19/24 questions) when asked on COVID-19 related knowledge. Knowledge data showed that the main source of information were healthcare workers, TV, friends and radio. The results indicate a significant number of healthcare workers did have sufficient knowledge on COVID-19 prevention.

This proportion of healthcare workers with adequate knowledge seems to conform with other studies in literature. A study done in Nepal reported a percentage of 76% while a study in Uganda found about 69% of healthcare workers with adequate knowledge^7,8^Another study conducted in Zambia among medical laboratory personnel reported that 84.1% of the medical laboratory workers had demonstrated sufficient knowledge on COVID-19.^9^ This is also reflected in the mean knowledge score for this study which was 80% only slightly lower than that of the Ugandan study which had a mean knowledge score of 82.4% among HCWs at MaKCHS Teaching Hospitals^9^ and equivalent to that found in a KAP toward COVID-19 among US residents (80%).^9,10^

This study revealed that knowledge level was especially influenced by level of education. People who had attained highest level of education were more likely to have higher knowledge scores compared to those who attained only secondary level of education. This is similar to the findings in a study conducted by which found that having a Bachelor’s degree was associated with an increase in the knowledge score among healthcare workers.^7^ Thus, further education and training through continuous professional education and journal clubs, particularly on symptoms and transmission would be essential in improving the knowledge of HCW about COVID-19 in our setting.^9^

This study also found that 60.3% of the healthcare workers had positive attitudes towards COVID-19. That is an average attitude score of 76% (15/20). Generally, this study is in agreement with studies conducted in Zambia among medical laboratory professionals and Vietnam which both found that majority of the healthcare workers had positive attitudes towards COVID-19 prevention.^9,11^ Findings in the study on the proportion of health workers with a positive attitude is consistent with what was found in another study were 55.7% of the healthcare workers had positive attitudes towards COVID-19 prevention.^8^

This study further revealed that age and level of education influenced attitudes towards COVID-19 prevention. Older people aged 25 – 29, 30 – 34, 35 – 39, 40 – 45 years were more likely to have positive attitudes compared to younger persons aged 20 – 24 years. Similarly, another study found that healthcare workers with a higher age were also more likely to have positive attitudes towards COVID-19.^12^ The finding in this and other studies can be explained by the fact that the higher the age, the longer the experience in dealing with emergencies, ultimately demonstrating confidence and optimism. Hence, increasing age could be the reason for a positive attitude.^13^

Additionally, sufficient knowledge on COVID-19 prevention was associated with a positive attitude. This is similar to the other findings by where it was found that there was a significant association between knowledge, attitude and practice at the level of *p* = 0.01^8^. Similarly, another study reported that there was a relationship between attitude score and knowledge score.^14^ An increase in knowledge was associated with an increase in the attitude score. This observation is equally reported in the study conducted by which reported that attitudes score was significantly associated with practices score (*p* = 0.009) indicating that subjects with a higher attitudes score are more likely to perform practices towards the prevention of SARS-CoV-2 transmission.^15^

Furthermore, this study reports that 59.9% of the healthcare workers had demonstrated good practice of the COVID-19 measures among the healthcare workers. This finding conforms to others results on practice among healthcare workers recorded in Nepal which revealed that over 60% had appropriate practice. Similarly, a study in Zambia among medical laboratory professionals found that 75% had good practice regarding COVID-19.^7^

This study found that the various factors that are associated with practice were only knowledge and attitude after adjusting for marital status, training, highest level of education and profession.

Most participants with poor practices are those with poor knowledge scores, without prior COVID-19 training and those with negative attitudes. Poor practices can lead to delayed and/or wrong laboratory diagnosis leading to poor patient management or safety incidents that could harm the personnel and their immediate coworkers, families and patients or healthcare workers.^16^

On the other hand, literature also provides evidence of findings that attitude and knowledge are two important predictors of COVID-19 related practices, for instance some literature study by reports that knowledge significantly influenced practice (*p* = 0.029)^17^ It further mentions that lack of personal protective equipment, fear of dying and going to common places, had a significant impact on the attitude of workers. The results in this study revealed that most of the healthcare workers did not have access to the adequate personal protective equipment and others indicated that some of the measures such as physical distancing were difficult to apply in the hospital set up. The shortage of PPEs, such as facemask, face shield, gloves, goggles, and gown, during this COVID-19 crisis are the major problems faced by not only the developing countries like Zambia, but also the developed world like the USA, UK, and Italy. Being a developing country, an adequate supply of PPEs is a significant challenge in Zambia and provides an explanation as to why this challenge was most cited in this study by the study participants. Ultimately, better knowledge may result in positive perceptions and attitudes and therefore in good practices, thus aiding in the prevention and management of infectious diseases, especially when the skills and resources are provided.^15^

### Conclusion

The focus of the study was established to assess the Knowledge, Attitude and Practice among healthcare workers towards Covid-19 preventive measures in Women and Newborn Hospital of the University Teaching Hospital. The study revealed that majority of the healthcare workers had adequate knowledge on COVID-19 related prevention, positive attitudes towards COVID-19 prevention and appropriate practices towards COVID-19 prevention. The study further revealed that individuals who had been to college or university were more likely to have sufficient knowledge. Sufficient knowledge was also associated with a positive attitude towards COVID-19 measures while knowledge and attitude were significantly associated with the practice of COVID-19 measures.

### Recommendations

Future studies to look at mixed method approach where both qualitative and quantitative data can be collected for heightened knowledge and validity.

### Ethical considerations

Approval was obtained from the University of Lusaka; the Women and Newborn Hospital. Additionally, approval was obtained from the National Health Research Authority. All participants in the study were required to participate voluntarily by providing informed consent, they were assured that no harm would come upon them should they decide not to take part in the study and they were not required to provide their personal identification information.

## Supporting information

strobe check list

## Data Availability

All data produced in the present study are available upon reasonable request to the authors

## AUTHOR CONTRIBUTIONS

JNS

- Work conception. Data acquisition, analysis and interpretation.
- Manuscript drafting.
- Approval of final manuscript.
- Accountable for all aspects of the work regarding its accuracy or integrity.

GL

- Work conception, Data analysis and interpretation.
- Manuscript drafting.
- Approval of final manuscript.
- Accountable for all aspects of the work regarding its accuracy or integrity.

MZ

- Manuscript drafting.
- Critical revisions for intellectual content.
- Data interpretation
- Approval of final manuscript.
- Accountable for all aspects of the work regarding its accuracy or integrity.

PJC

- Manuscript drafting.
- Critical revisions for intellectual content.
- Data interpretation
- Approval of final manuscript.
- Accountable for all aspects of the work regarding its accuracy or integrity.

## Acknowledgement

This article is a part of the master thesis submitted to the University of Lusaka in partial fulfilment of the requirements for the degree of Master of Public Health. The authors would like to thank the Ministry of Health and management at Women and Newborn Hospital for allowing us to use their institution to conduct our research.

